# A blood miRNA signature associates with sporadic Creutzfeldt-Jakob disease diagnosis

**DOI:** 10.1101/2020.01.08.19015214

**Authors:** Penny J. Norsworthy, Andrew G.B. Thompson, Tze H. Mok, Fernando Guntoro, Luke C. Dabin, Akin Nihat, Ross W. Paterson, Jonathan M. Schott, John Collinge, Simon Mead, Emmanuelle A. Viré

## Abstract

Sporadic Creutzfeldt-Jakob disease (sCJD) presents as a rapidly progressive dementia which is usually fatal within six months. No clinical blood tests are currently available for diagnosis or disease monitoring. Here, we profiled blood microRNA (miRNA) expression in sCJD. Small RNA-sequencing of 57 sCJD patients and 48 healthy controls revealed differential expression of hsa-let-7i-5p, hsa-miR-16-5p, hsa-miR-93-5p and hsa-miR-106b-3p. Downregulation of hsa-let-7i-5p, hsa-miR-16-5p and hsa-miR-93-5p replicated in an independent cohort using quantitative PCR, with concomitant upregulation of four of their mRNA targets. Absence of correlation in cross-sectional analysis with clinical phenotypes paralleled the lack of association between rate of decline in miRNA expression and rate of disease progression in a longitudinal cohort of 50 samples from 21 sCJD patients. Finally, the miRNA signature showed a high level of accuracy in discriminating sCJD from Alzheimer’s disease (AD). These findings highlight novel molecular alterations in the periphery in sCJD which can provide information about differential diagnosis and improve mechanistic understanding of human prion diseases.

## Introduction

Sporadic Creutzfeldt-Jakob disease (sCJD) is a transmissible neurodegenerative condition which typically develops in late middle age, is currently untreatable, and usually results in death within six months of symptom onset. Its characteristic pathological features are the development of spongiform change in the cerebral grey matter associated with deposition of abnormal forms of prion protein (PrP)^1^. sCJD is thought to be caused by the spontaneous formation of prions, an infectious agent comprised of multimeric assemblies of misfolded PrP^2,3^. Susceptibility to, and progression of, the disease is known to be strongly influenced by a common polymorphism at codon 129 of the prion protein gene (PRNP)^4,5^, and additionally, several different strains of misfolded PrP are known to exist which can influence the course of the disease^6^. Prions are highly concentrated in brain tissue at post mortem examination of sCJD, but are also detectable in peripheral tissues, with no apparent biological consequences^7-10^. Specific diagnostic tests are available including brain magnetic resonance imaging (MRI) and cerebrospinal fluid (CSF) analyses; however, no simple blood assay has been developed. Here we investigated blood microRNA (miRNA) profiles to explore whether these would disclose novel accessible biomarkers or insights into the peripheral pathobiology of the most common human prion disease.

Small non-coding RNAs, such as miRNAs, are highly conserved oligonucleotides (19-22 bases) which can inhibit translation of targeted messenger RNA (mRNA) transcripts and tag them for degradation^11^. miRNAs are powerful regulators of gene expression because one miRNA can target multiple coding genes. Due to their remarkable stability in body fluids^12^, miRNAs represent attractive biomarkers and drug targets, and their dysregulation has been widely described in disease states^13-15^. Moreover, since miRNAs in blood often originate from other tissues, the study of miRNAs can provide novel insights into pathological mechanisms in inaccessible tissues such as the brain^12^. miRNAs have been extensively studied in Alzheimer’s disease (AD), Parkinson’s disease (PD), Huntington’s disease (HD) and Amyotrophic Lateral Sclerosis (ALS)^16-19^.

In prion disease, only a few studies have investigated the contribution of miRNAs to disease initiation and progression, and most of the work has focused on animal models and brain tissue, limiting existing knowledge in humans to the end-stage of the disease^20^. Recently, Slota et al profiled serum miRNAs in chronic wasting disease, a prion disease of deer^21^, demonstrating their potential as biomarkers in animals. Here, we profiled miRNA expression levels in whole blood by case-control study. Small RNA-sequencing (RNA-seq) identified four differentially expressed (DE) miRNAs. Using quantitative PCR (qPCR) in an independent cohort we validated and replicated the finding that hsa-let-7i-5p, hsa-mir-16-5p and hsa-mir-93-5p are downregulated in sCJD. To gain further insight into the potential use of the signature in differential diagnosis, we measured levels of these miRNAs in whole blood from patients with AD and showed that the miRNA signature in sCJD is clearly differentiated from AD. We also observed that expression of downstream mRNA targets of these miRNAs are affected in sCJD blood. Further, by interrogating 50 longitudinal samples from a group of 21 sCJD patients we showed that the rate of change of DE miRNA expression did not correlate with the rate of disease progression.

## Results

### Sequencing of small RNA in blood identifies five miRNAs differentially expressed between sCJD cases and healthy controls

To investigate if miRNA profiles could discriminate patients with sCJD from healthy controls, we designed a case-control study using blood from 57 sCJD cases and 48 controls (Table 1). Using next-generation sequencing, we measured expression of miRNAs in all samples. Across all sequencing runs, the mean percentage of clusters passing filter was 84.43%, and the mean percentage of bases with Q>30 was 84.80%. Across all samples, 94.73% of reads were successfully aligned to the genome, and 94.72% of these reads aligned uniquely. The mean Q score for aligned reads was 36.6. After alignment, quantification, and normalisation of reads, 866 unique miRNAs were detected. To represent the overall expression pattern of the miRNAs in all 105 samples, we performed principal component analysis (PCA) (Supplementary Fig. 1). 101 miRNAs which passed low expression filtering were analyzed for differential expression (Supplementary Data 1). Figure 1a, and Supplementary Data 1, show that four miRNAs were found to have a statistically significant fold change (FC) between cases and controls after False Discovery Rate (FDR) correction for multiple testing. Figure 1b shows that hsa-miR16-5p (FC –2.76, adjusted p=1.48×10”^4^), hsa-miR-93-5p (FC –2.34, adjusted p=0.001), hsa-miR-106b-3p (FC –1.74, adjusted p=0.008), and hsa-let7i-5p (FC –2.49, adjusted p=0.011), were downregulated in sCJD compared to healthy controls. hsa-let-7d-3p was also included because its levels were elevated in sCJD blood, albeit with borderline significance (FC 1.94, adjusted p=0.053). Age was included as a covariate in the regression test, although none of these five miRNAs showed a significant correlation with age in controls. Because sCJD does not alter blood cell counts^22^, normalisation of the sequencing data using cell count data was not done. Although the RNA we were able to obtain for a proportion of sCJD patients in our study was sub-optimal in terms of RNA Integrity Number (RIN) for mRNA analysis, miRNA is highly resistant to degradation^23^, and an independent analysis of the small RNA-seq data, excluding RNA samples where RIN<4, did not alter the findings reported here (Supplementary Table 1). Because a small number of the sCJD cases had a ‘probable’ diagnosis rather than definite (Table 1), we also carried out a third analysis in which we excluded these three ‘probable’ patients (Supplementary Table 2). This did not affect the findings. Finally, 36 samples (14 sCJD cases and 22 controls) yielded more than 1.5 million reads. Given that a small number of miRNAs make up the majority of read counts in blood, we analysed these 36 samples in order to take lower abundance miRNAs into account. This added another 30 miRNAs to the list of those tested for differential expression between sCJD and controls. None of these low abundance miRNAs were significantly differentially expressed in this analysis (Supplementary Data 2). We therefore only followed up the five above mentioned miRNAs in our downstream analyses.

**Table 1.** Patient data by study group.

|  | Discovery Cohort |  | Replication Cohort |  | AD Cohort |  | Longitudinal sCJD cohort (n=21) |  |
| --- | --- | --- | --- | --- | --- | --- | --- | --- |
| Group | sCJD | Control | sCJD | Control | AD | Control | No. samples/patient, median (IQR<sup>a</sup>) | 2 (2-3) |
| n | 57 | 48 | 29 | 30 | 30 | 30 | Sampling interval, median (IQR), days | 52 (29-92) |
| Female/Male, n (%) | 31/26 (54/46) | 29/19 (60/40) | 12/17 (41/59) | 13/17 (43/57) | 12/18 (40/60) | 13/17 (43/57) | Female/Male, n (%) | 8/13 (38/62) |
| Sampling age, mean (SD<sup>b</sup>), yrs | 66 (8) | 54 (15) | 63 (7) | 59 (13) | 61 (5) | 59 (13) | Age at first sample, mean (SD), yrs | 63 (8) |
| RIN<sup>c</sup>, mean (SD) | 5.6 (1.3) | 6.5 (1.2) | 5.8 (1.8) | 6.8 (1.0) | 6.6 (0.6) | 6.8 (1.0) | RIN (all samples), mean (SD) | 5.9 (1.2) |
| Definite/Probable sCJD, n (%) | 54/3 (95/5) | N/A | 28/1 (97/3) | N/A | N/A | N/A | Definite/Probable sCJD, n (%) | 13/8 (62/38) |
| *PRNP* codon 129 genotype<sup>d</sup>, n (%) | 26/15/16 (46/26/28) | 16/24/8 (33/50/17) | 10/12/6 (36/43/21) | 7/7/5 (37/37/26) | N/A | N/A | *PRNP* codon 129 genotype, n (%) | 3/11/7 (14/53/33) |
| MRC Scale score<sup>e,54</sup>, mean (SD) | 6.86 (5.48) | 20 (0) | 7.89 (5.81) | 20 (0) | N/A | N/A | MRC Scale slope<sup>f,4</sup>, median (IQR) | 0.353 (0.185-0.823) |

<sup>a</sup>Interquartile Range; <sup>b</sup>Standard Deviation; <sup>c</sup>RNA Integrity Number; <sup>d</sup>In order MM/MV/VV, where numbers do not add up to group totals this is due to unavailability of a small number of samples; <sup>e</sup>MRC Prion Disease Rating Scale score, a measure of disease severity from 0-20, with 20 being unaffected;
<sup>f</sup>Rate of disease progression measured in % reduction in function per day; N/A Parameter not relevant.

**Fig.1.**
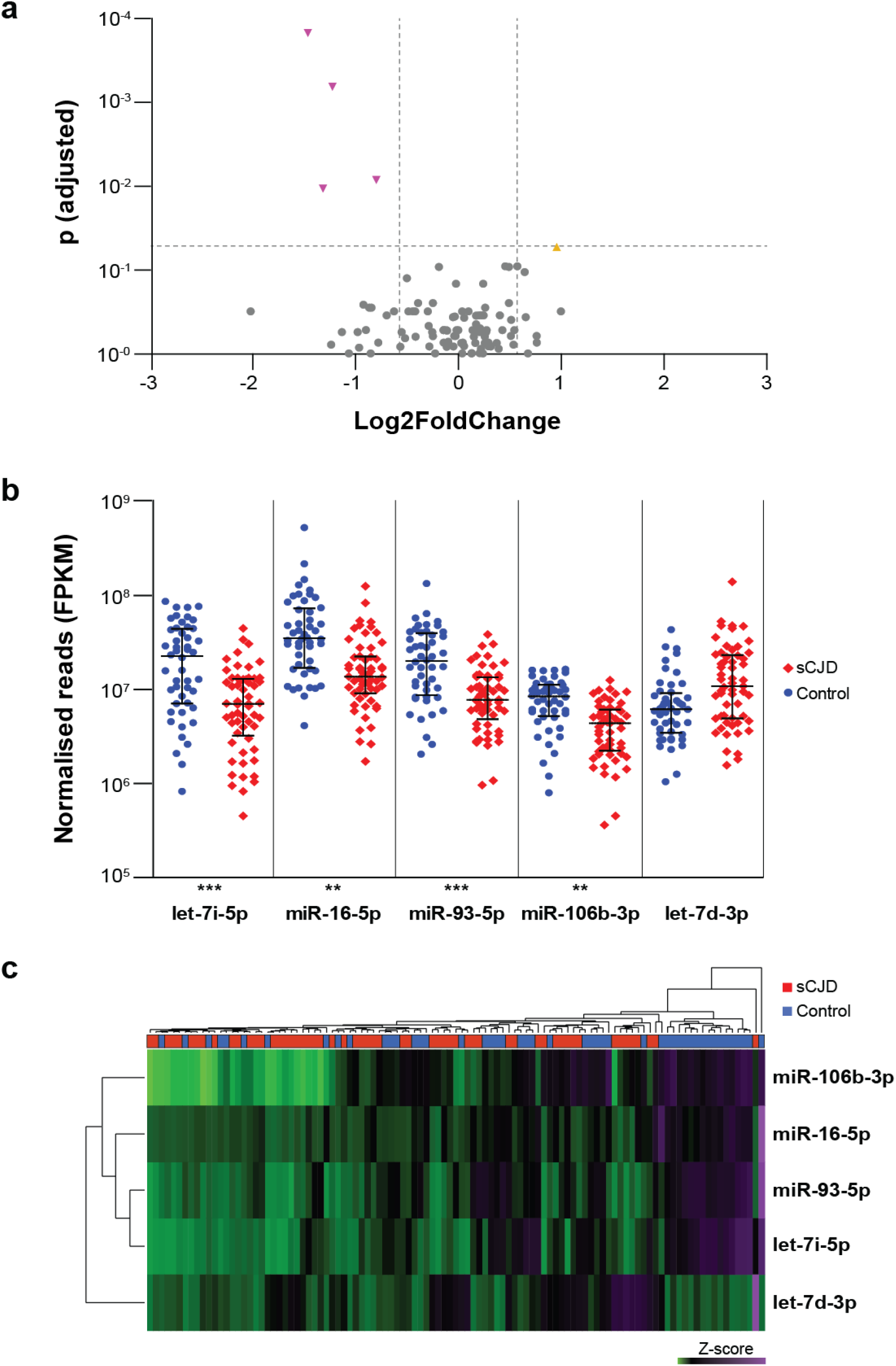
Differential expression of miRNAs in the discovery cohort measured by small RNA-seq. Volcano plot of 101 miRNAs analyzed for differential expression (**a**): vertical lines are shown to delineate >1.5 fold change; horizontal line to delineate adjusted p<0.05. miRNAs followed up for validation and replication are shown by the yellow triangle (upregulated) and upside-down triangles (downregulated).(**b**), plotted by normalised transcript count (in Fragments Per Kilobase of transcript per Million mapped reads, FPKM) with median and interquartile range are shown for both control and sCJD groups. hsa-miR-let-7d-3p showed borderline significance at p = 0.053. **p<0.01, ***p<0.001. Control (blue circles) n=48; sCJD (red diamonds) n=57. Hierarchical clustering of DE miRNA profiles (**c**), illustrating relationships between both DE miRNAs, and individuals within the discovery cohort. Control (blue) n=48; sCJD (red) n=57. Source data for (**a**) and (**b**) are provided as a Source Data file.

Unsupervised hierarchical clustering analysis using these five differentially expressed miRNAs yielded two major clusters of samples, with most healthy individuals clustering tightly together (Fig. 1c). When the data was stratified based on *PRNP* codon 129 (either MM, MV, or VV) (Supplementary Fig. 2) or PrP^Sc^ strain type (the prion protein disease isoform typed by the London classification, types 2 or 3, n=22), no significant differences were observed, suggesting that these factors have little or no effect on expression of these miRNAs.

In a subset of the 866 detected miRNAs with sufficient data for analysis (including those excluded from differential expression analysis by the low expression filter), expression in sCJD patients was tested for correlation with clinical parameters (age of onset, duration of disease, MRC Scale score and MRC Scale slope). No significant associations were observed after correction for multiple testing.

### Signature of differentially expressed miRNAs in blood from sCJD patients is replicated in an independent cohort

Having identified a blood-based miRNA signature for sCJD, we aimed to validate and replicate these results using an independent cohort and technique. The replication cohort comprised 29 sCJD patients and 30 healthy age and sex matched control individuals (Table 1). Reverse transcription quantitative PCR (RT-qPCR) was used to measure the relative levels of expression of the five differentially expressed miRNAs. hsa-miR-484, which was not differentially expressed in the discovery study (FC 1.07, adjusted p=0.658), was also included. Having determined that levels of endogenous controls did not vary between cases and controls (Supplementary Fig. 3d), we used RNU6-2, SNORD42B, SNORD95 and SNORD72 to normalise miRNA expression levels. Lower levels of hsa-let-7i-5p, hsa-miR-16-5p and hsa-miR-93-5p were found in sCJD blood, whilst hsa-miR-484 remained unchanged as expected (Fig. 2a, and Supplementary Fig. 3a-c), thereby replicating the findings from the discovery study. Of the remaining two miRNAs tested, hsa-miR-106b-3p amplified poorly, while hsa-let-7d-3p was not replicated (Fig. 2a, and Supplementary Fig. 3a-c).

**Fig.2.**
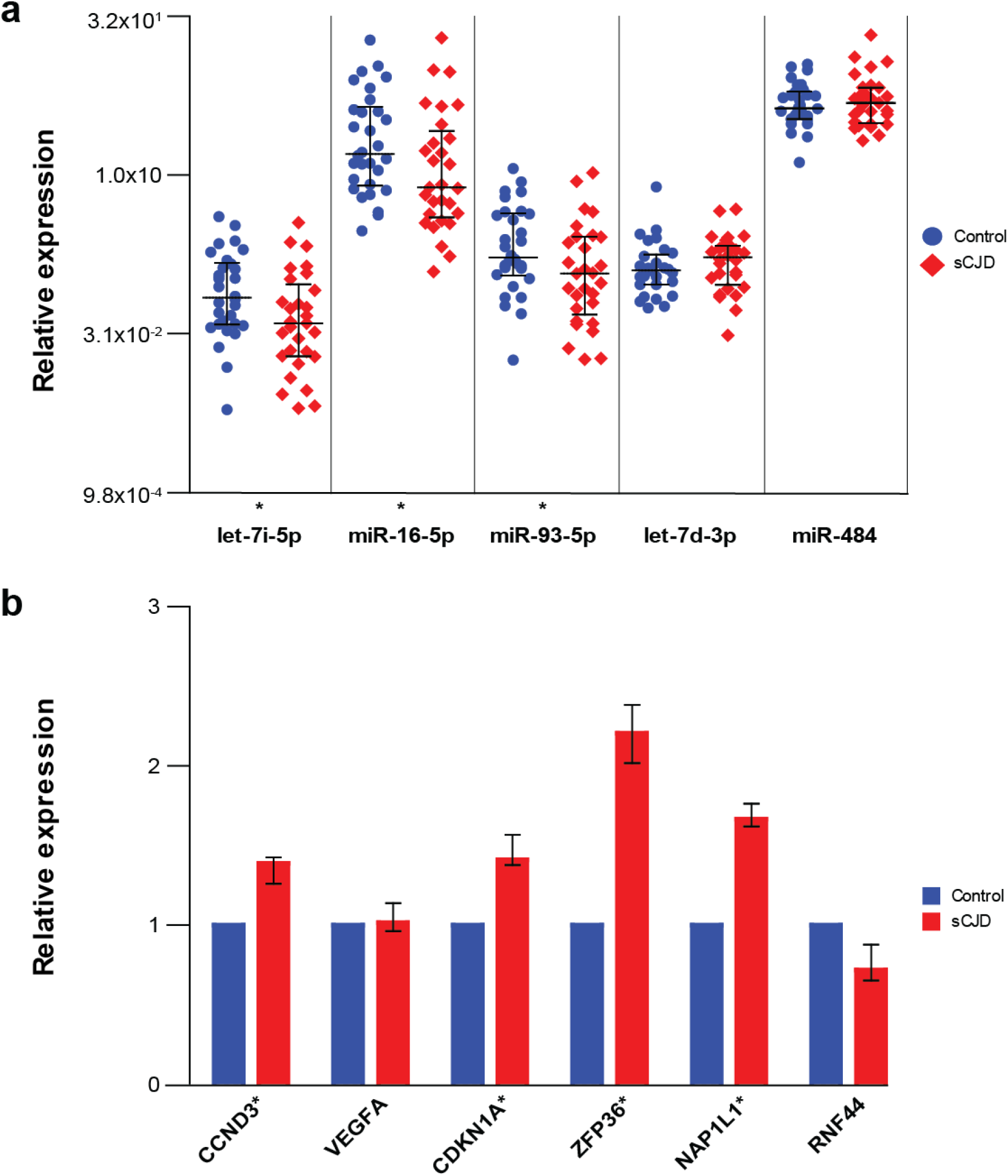
Validation and replication of DE miRNAs, and expression of target mRNAs measured by qRT-PCR. Expression of DE miRNAs (**a**), relative to the control small RNA RNU6-2 with median and interquartile range, are shown for both control and sCJD groups. Fold change (FC) as follows: hsa-let-7i-5p FC - 1.82; hsa-miR-16-5p FC -1.87; hsa-miR-93-5p FC -1.74; hsa-let-7d-3p FC 1.14; hsa-miR-484 FC 1.09; *p<0.05. Control (blue circles) n=30; sCJD (red diamonds) n=29. Expression of target mRNAs (**b**), plotted relative to combined expression of control mRNAs *(ALAS1, B2M* and *ACTB)* in control and sCJD groups. Median and range are shown. Median FC as follows: *CCND3* FC 1.39; *VEGFA* FC 1.02; *CDKN1A* FC1.41; *ZFP36* FC 2.21; *NAPL1L* FC 1.67; *RNF44* FC -1.43. *p <0.05. Control (blue) n=30; sCJD (red) n=29. Source data for (**a**) and (**b**) are provided as a Source Data file.

### miRNA targets are altered in blood from sCJD patients

Next, we hypothesized that if differential miRNA expression plays a role in the peripheral pathophysiology of sCJD, relevant mRNA targets should also be altered in sCJD blood. We identified these mRNA targets using TarBase with stringent criteria (validated using luciferase assays and/or validated by immunoprecipitation; expressed in whole blood) (Supplementary Table 3). Using RT-qPCR we measured the expression of six transcripts reported to be under the control of hsa-let-7i-5p, hsa-miR-16-5p and hsa-miR-93-5p in the replication cohort: *CCND3, VEGFA, NAP1L1, ZFP36, CDKN1A* and *RNF44*. Figure 2b shows that blood samples from sCJD patients display higher levels of four of these transcripts in comparison to blood from controls, which is consistent with a reduction of miRNA silencing of target mRNA. Mean endogenous control Ct values showed no significant differences between control and sCJD groups (Supplementary Fig.4). Finally, to gain insight into biological processes by which these miRNAs may contribute to sCJD, we performed pathway enrichment analysis using the six targets of the Differentially Expressed (DE) miRNAs and found no significant enrichment. Altogether, these results demonstrate that hsa-let-7i-5p, hsa-miR-16-5p and hsa-miR-93-5p are downregulated in sCJD blood, and concomitantly, levels of their selected mRNA targets are elevated.

### Downregulation of hsa-let-7i-5p, hsa-miR-16-5p and hsa-miR-93-5p is specific to sCJD

To further investigate the relevance of this blood-based miRNA signature to sCJD we next set out to profile levels of hsa-let-7i-5p, hsa-miR-16-5p and hsa-miR-93-5p in blood collected from the first specialist assessment of patients with AD (equivalent to sampling at the first opportunity in sCJD patients). A group of 30 AD patients were matched by sex and age to the control group used for the replication analysis (Table 1). RT-qPCR data presented in Figure 3 and Supplementary Fig. 5a-c, show that, in marked contrast to sCJD, hsa-let-7i-5p, hsa-miR-16-5p and hsa-miR-93-5p were upregulated in AD patients. Mean endogenous control Ct values showed no significant differences between control and AD groups (Supplementary Fig. 5d). These results demonstrate that levels of hsa-let-7i-5p, hsa-miR-16-5p and hsa-miR-93-5p in blood establish a signature which enables discrimination between healthy controls, AD, and sCJD.

**Fig.3.**
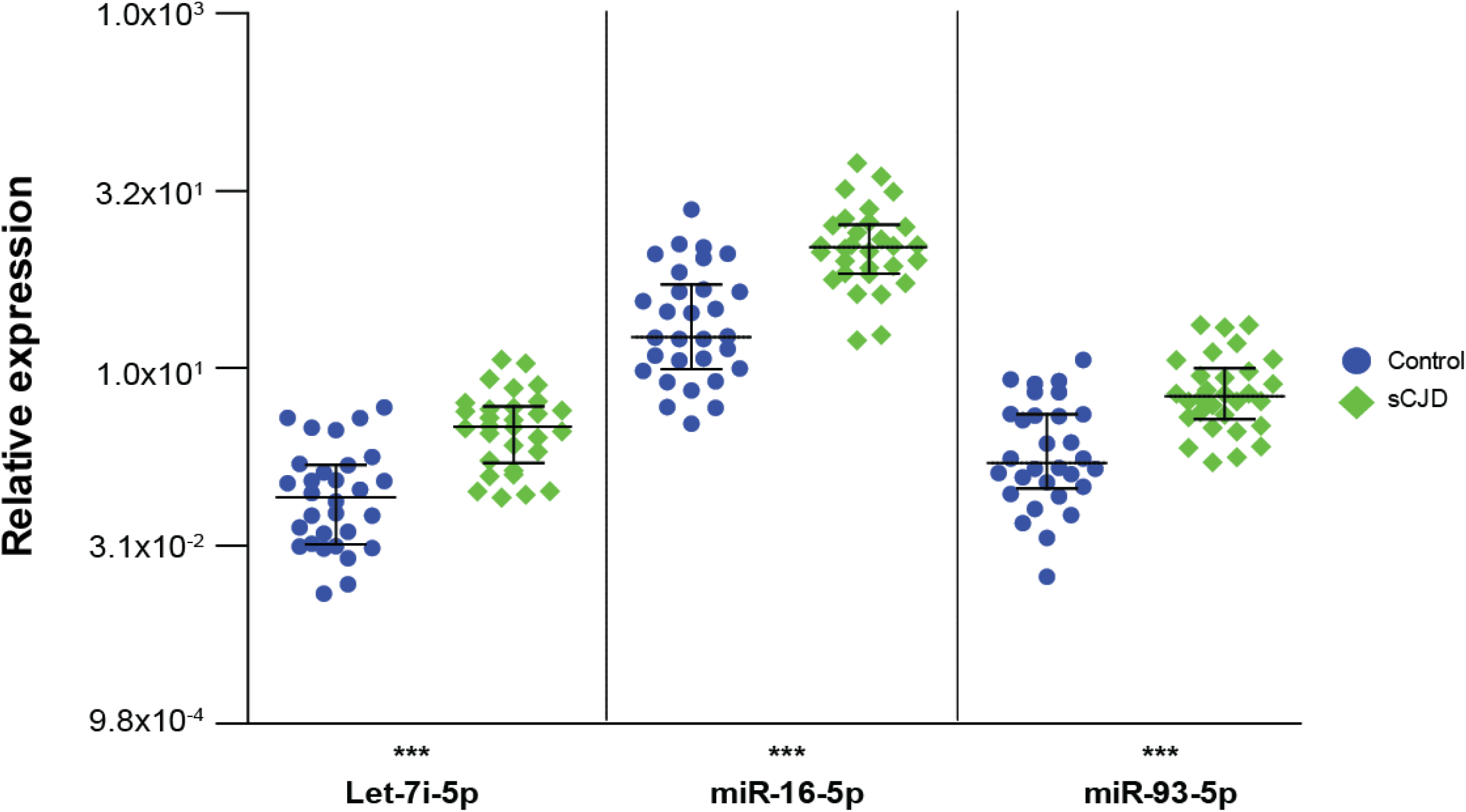
Expression of replicated DE miRNAs in Alzheimer’s Disease (AD) measured by qRT-PCR. Expression relative to the control RNA RNU6-2 with median and interquartile range are shown for control and AD groups. hsa-let-7i-5p FC 3.84; hsa-miR-16-5p FC 4.46; hsa-miR-93-5p FC 3.22. ***p<0.001. Control (blue circles) n=30; AD (green diamonds) n=30. Source data are provided as a Source Data file.

### Rate of decline in miRNA levels does not correlate with sCJD rate of progression

Although there was no cross-sectional correlation between disease metrics and DE miRNAs, we asked whether the rate of decrease in miRNA expression could be related to the speed of decline observed in individual patients with sCJD. High inter-individual variation might obscure intra-individual longitudinal effects over time. To this end, 50 bloods were collected longitudinally from 21 individuals with sCJD (Table 1 and Supplementary Data 3). At time of sampling, each patient’s condition was assessed using the MRC Prion Disease Rating Scale, allowing for disease progression to be monitored (% loss of function per day by mixed effects regression). Levels of expression for hsa-let-7i-5p, hsa-miR-16-5p, hsa-miR-93-5p and hsa-miR-484 (which was found to be unaltered in sCJD blood) were quantified by RT-qPCR and values were converted into rate of expression decline (reduction in expression per day). Figure 4a-d and Supplementary Fig. 6-7 shows that rate of decline in levels of hsa-let-7i-5p, hsa-miR-16-5p and hsa-miR-93-5p did not correlate with rate of disease progression (% loss of function per day).

**Fig.4.**
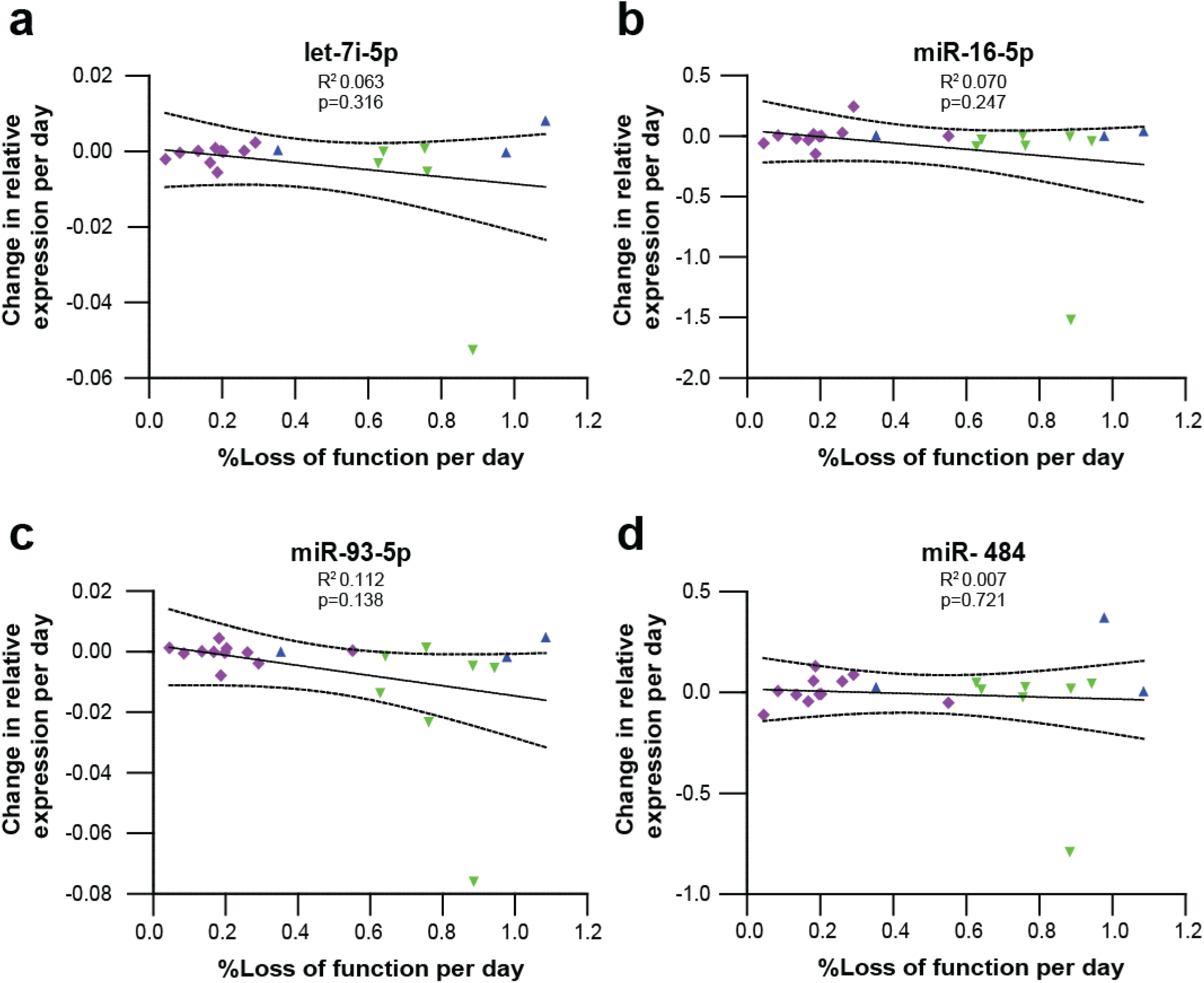
Relationship between disease progression and change in DE miRNA levels in longitudinal samples from sCJD patients. miRNA expression was measured relative to the control small RNA RNU6-2 for (**a**) hsa-let-7i-5p, (**b**) hsa-miR-16-5p, (**c**) hsa-miR-93-5p and (**d**) hsa-miR-484. hsa-miR-484 is a non-DE miRNA shown here for comparison. %Loss of function per day was calculated from decline in MRC Scale score. R^2^ and p values are shown. Dotted lines represent 95% confidence intervals. Data points are coded by *PRNP* codon 129 genotype (MM blue triangle, MV magenta diamonds, VV green inverted triangle). n=21 (18 for let-7i-5p). Source data for (**a-d**) are provided as a Source Data file.

### Diagnostic potential of miRNA markers

Finally, to assess the capabilities of miRNAs to discriminate between sCJD cases and healthy controls, we applied Receiver Operating Characteristic (ROC) analysis. Using the data from the discovery study, the normalised read count of each of the three replicated DE miRNAs was log transformed to ensure normal distribution, and Z scores were calculated (distance from the mean in SDs) across the 105 samples in the discovery study. This analysis was performed for each miRNA individually, and for the aggregated mean of the three sCJD specific miRNAs (hsa-let-7i-5p, hsa-miR-16-5p, hsa-miR-93-5p). In the discovery dataset, comparing sCJD to healthy controls, the Area Under the Curves (AUCs) ranged from 0.736 (has-let-7i-5p) to 0.762 (hsa-miR-16-5p) and reached 0.788 when all three miRNAs were combined (Fig. 5a and Supplementary Table 4). In the replication set, all three miRNAs aggregated performed better than each of them individually (Fig. 5b and Supplementary Table 4). Because we found that hsa-let-7i-5p, hsa-miR-16-5p and hsa-miR-93-5p were upregulated in AD patients (as opposed to downregulated in sCJD), we next performed the same analysis (i.e. comparing AD to healthy controls) using the three miRNAs individually and aggregated. Supplementary Table 4 and Figure 5c show that AUC values ranged from 0.810 (hsa-miR-93-5p) to 0.859 (hsa-miR-16-5p) and reached 0.860 for the three miRNAs together, indicating good diagnostic discrimination. Finally, given the lack of biomarkers to specifically and selectively discriminate sCJD patients from AD patients in blood, we calculated AUCs for the sCJD versus AD comparisons. Strikingly, AUC values ranged from 0.897 (hsa-miR-93-5p) to 0.934 (hsa-let-7i-5p). Combining the 3 miRNAs together yielded AUC value of 0.924 and 100% specificity (Fig. 5d and Supplementary Table 4). These results were not only seen when normalising to RNU6-2. Similar findings were made using SNORD42B, SNORD95 and SNORD72 endogenous controls (Supplementary Fig. 8-10). Altogether, these results suggest that blood miRNA expression markers can be used to discriminate AD from healthy controls and sCJD from AD.

**Fig. 5.**
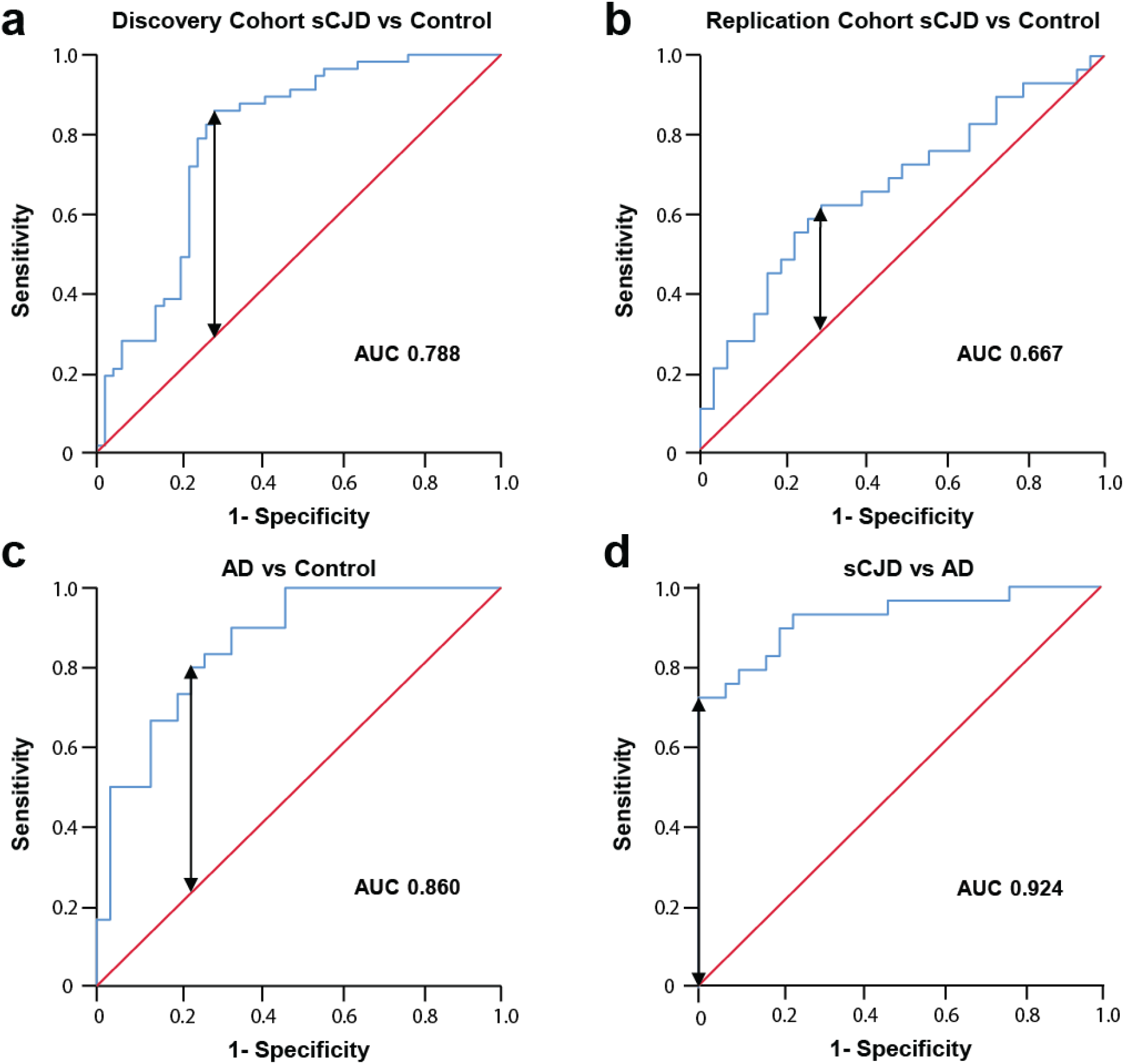
Diagnostic potential of blood miRNA biomarkers. ROC curves were constructed using mean Z scores calculated from (**a**) log transformed normalised small RNA-seq reads in sCJD versus control individuals in the discovery study. sCJD n=57; control n=48; and (**b**) Ratios of hsa-let-7i-5p, hsa-miR-16-5p and hsa-miR-93-5p measured relative to RNU6-2; sCJD versus control individuals in the replication study. sCJD n=29; control n=30; (**c**) Ratios of hsa-let-7i-5p, hsa-miR-16-5p and hsa-miR-93-5p measured relative to RNU6-2; AD versus control individuals. AD n=30; control n=30; and (**d**) Ratios of hsa-let-7i-5p, hsa-miR-16-5p and hsa-miR-93-5p measured relative to RNU6-2; sCJD versus AD. sCJD n=29; AD n=30. Area under the curve (AUC) is shown for each comparison; maximal Youden’s Index is indicated by the double headed arrow, where test performance is optimal. The line of zero discrimination is shown in red. Source data for (**a-d**) are provided as a Source Data file.

## Discussion

Sporadic Creutzfeldt-Jakob disease, the most common human prion disease and the archetypal rapidly progressive dementia, presents with neurological and/or neuropsychiatric features, and diagnosis is supported by abnormalities of brain imaging, neurophysiology or CSF analysis. Currently available blood tests do not help to distinguish sCJD from other neurodegenerative disorders. Here, for the first time, we show replicated evidence of specific dysregulation of miRNAs in the peripheral blood of sCJD patients. Using next-generation sequencing to profile miRNA expression in peripheral blood, we identified, validated, and replicated three miRNAs (hsa-let-7i-5p, hsa-miR-16-5p and hsa-miR-93-5p) whose expression is significantly lower in the blood of sCJD patients than in healthy individuals. Furthermore, we report that the levels of these three miRNAs distinguish patients with sCJD from patients with AD. These findings suggest there is a peripheral blood molecular signature in sCJD that could be explored for potential future applications as a biomarker for diagnosis.

Each dysregulated miRNA could plausibly have implications for the pathobiology of sCJD. hsa-miR-93-5p is a member of the miR-106b-25 miRNA cluster (comprising hsa-miR-106b, hsa-miR-93 and hsa-miR-25), located on chromosome 7 within the *MCM7* gene. Expression of both the miR-106b-25 cluster and *MCM7* (which is also downregulated in sCJD blood, Supplementary Fig. 11) is regulated by the transcription factor E2F1, a cell cycle regulator which has been associated with neurotoxicity in neurodegenerative disease^24-26^. In the discovery cohort the other two members of the cluster, hsa-miR-106b-3p and hsa-miR-25-3p, were also downregulated in sCJD patients. However, expression of hsa-miR-106b-3p was too low to allow replication by qPCR, and the adjusted p value for hsa-miR-25-3p fell short of significance (p=0.125, Supplementary Data 1). Repression of this cluster has previously been shown to be required for endoplasmic reticulum (ER) stress induced apoptosis, a common feature of neurodegenerative diseases^27,28^. It is unsurprising to find that members of this cluster are dysregulated in sCJD since alterations in hsa-miR-93-5p expression have been previously linked to AD and ALS^18,29^, and the prion protein gene, *PRNP*, has been identified as a potential target of hsa-miR-93-5p^30,32^.

hsa-let-7i-5p has also been implicated in neurodegenerative disorders. In ALS, Liguori *et al* reported downregulation of hsa-let-7i-5p in whole blood^18^. Conversely, in AD, elevated levels of hsa-let-7i-5p were found in brain, plasma and CSF^33,34^. These findings are consistent with our observations of hsa-let-7i-5p’s downregulation in sCJD blood but upregulation in AD whole blood (Fig. 2a and 3). More relevant is the report that levels of hsa-let-7i-5p were found to be elevated in exosomes from neurons of prion-infected mice^35^, and in sera from elk with chronic wasting disease^21^

hsa-miR-16-5p features prominently in existing literature on miRNA and neurodegeneration, and prion disease in particular. Studies in prion-infected mice have shown a distinct temporal pattern of dysregulation of hsa-miR-16-5p as disease progresses, whereby expression is upregulated in cultured primary neurons in the pre-symptomatic phase but downregulated in the clinically established disease phase^36,37^. Recently, Slota et al. reported decreased miR-16-5p abundance in serum from scrapie infected hamsters^21^. Our observations of downregulation of hsa-miR-16-5p in blood from patients with clinically established sCJD concur with these findings. In humans, Llorens and colleagues reported increased levels of hsa-miR-16-5p in sCJD frontal cortex^38^. In ALS, hsa-miR-16-5p was found to be downregulated in whole blood^18^, and in AD, several clinical and mechanistic studies have suggested that hsa-miR-16-5p plays a role in pathogenesis^39-41^. Whether altered abundance in the blood is related to the mechanism by which it is differentially expressed in neurons or in neurodegeneration remains to be investigated.

The question of whether dysregulated miRNAs are shared between neurodegenerative disorders is crucial because of the unmet need for the treatment of these diseases. Therefore, lifting the veil on molecular mechanisms involved in neurodegeneration could help provide evidence for better disease classification and drug design. Together with the literature, our results suggest that the same small group of miRNAs is consistently altered in neurodegenerative disorders, with effect size and direction of effect being tissue and disease-stage specific. miRNA profiling may, in the future, help with disease classification and diagnosis for several reasons. Firstly, because our signature allows us to clearly distinguish sCJD from AD using whole blood: the decrease in hsa-let-7i-5p, hsa-miR-16-5p and hsa-miR-93-5p in sCJD blood contrasts markedly with their upregulation in AD blood (Fig. 2a and 3), and also AD serum, CSF and brain tissue ^33,34,42^. Secondly, because the profiles of dysregulated miRNAs are highly variable between disease stages and disease models. Kanata *et al* recently conducted a comprehensive review of changes in miRNAs reported in mouse and primate models of prion disease^20^. The authors pointed out that the variability in direction of effects reported could be caused by the wide variety of models studied, technologies used and tissues analyzed (e.g. post mortem brain versus whole blood from living patients), as well as disease state (pre-clinical and clinical)^37,38,43^. Consistent with this, the sCJD miRNA signature we identified here is also found in sporadic ALS when the same tissue is analyzed^18^. There are significant parallels between the two conditions in terms of pathology associated with deposits of aggregated misfolded protein, and heterogeneous clinical phenotypes^44-46^. The finding that dysregulated miRNAs are shared between these two diseases suggests that common mechanisms might be involved, but does not negate any potential diagnostic relevance of the sCJD signature, since the two diseases are clinically very different (ALS is predominantly a disorder of motor neurons, whilst sCJD presents with pervasive brain dysfunction). Also of note is the study from Llorens *et al*, whose findings demonstrated widespread dysregulation of miRNA levels in several brain regions affected by sCJD and implicated the miRNA biosynthesis pathway in sCJD pathogenesis^38^.

Insights into molecular mechanisms that can contribute to the disease process may also be provided by investigation of transcripts known to be targeted by the sCJD signature: the increased expression of several hsa-let-7i-5p, hsa-miR-16-5p and hsa-miR-93-5p target genes (Fig. 2b) demonstrates that the observed downregulation in these three miRNAs in sCJD patients has concomitant effects on target gene regulation. By performing analysis of DE miRNA target binding sequences, we excluded the possibility that the observed downregulation in miRNAs resulted from Target RNA-Directed MicroRNA Degradation (TDMD)^47^. Intriguingly *ZFP36* (in protein form known as tristetraprolin, TTP), which was the most highly overexpressed target gene we tested in sCJD compared with controls (Fig. 2b), is a stress granule associated RNA binding protein which has been linked with AD pathology in mice and humans^48,49^.

A major strength of this study was our ability to access longitudinal samples for 21 sCJD patients, which enabled us to test for association with disease progression. Absolute expression levels of the signature did not associate with disease severity, nor did the rate of decline of expression and rate of disease progression (Fig. 4a-d and Supplementary Fig. 6a-d and 7a-d). Whether these miRNAs are a driver or passenger of speed of decline in sCJD remains to be established. Although the use of patient samples is desirable, it does however pose multiple caveats and limitations: confounding factors associated with disease status such as sample handling and storage prior to extraction, RNA degradation, age, environmental exposure and interactions with drugs could lead to changes in miRNA expression profiles. We addressed concerns about RNA degradation by conducting a further analysis of the discovery cohort data to show that RIN number for total RNA did not have an impact on our findings (Supplementary Table 1). This is not surprising, as miRNA is known to be highly resistant to degradation in comparison to mRNA, and therefore RIN measured from total RNA does not reflect the degradation status of miRNA^23^. Another potential limitation of our study is that it uses next-generation sequencing, a technology based on direct counts. Less abundant miRNAs could be missed, therefore masking some disease specific patterns. Sequencing coverage in the discovery study is not high, but as hsa-miR-106b-3p could not be replicated by qPCR due to its low expression, inclusion of miRNAs expressed at lower levels by resequencing would have been unlikely to yield additional DE miRNAs that could be validated by qPCR.

Hemolysis is another potential confounding factor when profiling miRNA expression in patient blood samples^50^. This is because a number of miRNAs are known to be present at high levels in red blood cells, and their release could be masking disease related patterns. Here, we found that hsa-miR-484, whose levels are known to increase in hemolysed serum^51^, remained unchanged between sCJD and controls (both in sequencing and qPCR datasets). Our results are therefore unlikely to be caused by red blood cell contamination. Interestingly, Ludwig et al. recently showed that miRNAs with lower abundance in AD were enriched in monocytes and T-helper cells, while those up-regulated in AD were enriched in serum, exosomes, cytotoxic t-cells, and B-cells^52^.

To our knowledge, this study is the first to evaluate the use of blood miRNA expression as biomarkers in sCJD. This study revealed diagnostic potential of blood miRNAs, and particularly high accuracy, in discriminating AD patients from healthy controls, and sCJD patients from AD. Therefore, blood miRNA expression could potentially be used as a non-invasive marker to help differentiate between prion infection and other neurodegenerative diseases in humans. Future work will look at other differential diagnoses and help establish whether miRNA profiles used in combination with other biomarkers, and demographic and clinical features, improve diagnostic accuracy in neurodegenerative diseases.

Our work was conducted in whole blood which has the advantage of being an accessible tissue, which therefore enabled us to study a large group of patients from early disease stages, and longitudinally. It will be important to establish whether the findings reported here are replicated in other tissues and body fluids such as CSF and brain tissue in order to determine whether the miRNA signature we have identified in blood is present in the brain, with potential additional effects on brain specific mRNA targets. Based on GTEx datasets^53^, four of the mRNA targets of the three altered miRNAs are expressed in normal brain. Investigating the expression of both miRNAs and their targets in sCJD brain tissues will provide more evidence for a potential role in disease pathogenicity. Other future analyses might consider the impact of sex, ethnicity, co-morbidities, subtypes of disease and specific symptoms on the miRNA profile. In conclusion, our analysis of a large cohort of sCJD patients and controls has revealed, for the first time, that sCJD affects miRNA expression profiles in peripheral blood and that these changes are a signal in the periphery rather than just a consequence of brain degeneration. Additionally, we identified a three miRNA signature whose-expression can discriminate patients with AD from controls and sCJD from AD cases. These findings provide the basis for a better mechanistic understanding of sCJD and represent an opportunity to open much needed new avenues for improved disease diagnosis and therapeutic intervention.

## Methods

### Participants

Patients with a definite (i.e. neuropathologically confirmed) or probable diagnosis of sCJD (Table 1) according to World Health Organization criteria were recruited by the National Prion Clinic (London, UK). sCJD patients in the longitudinal cohort were participants in the UK National Prion Monitoring Cohort^54^. MRC Prion Disease Rating Scale scores (on a scale of 0-20, where 20 is unaffected) were derived for patients in the longitudinal cohort as previously described^54^, and rates of decline (% loss of function per day) calculated according to the linear mixed model method described in Mead *et al*^4^.

Healthy control donors were recruited from spouses or relatives of patients by the National Prion Clinic (London, UK), and the UCL Dementia Research Centre (London, UK). Patients diagnosed with AD according to International Working Group-2 criteria^55^ were recruited by the UCL Dementia Research Centre (London, UK) and other referrers in the UK from 2012-2018. The study was approved by the London – Harrow Research Ethics Committee (reference 05/Q0505/113). Samples were obtained with written informed consent from controls, patients, or a patient’s consultee in accordance with applicable UK legislation and Codes of Practice.

### Sample collection and RNA extraction

Whole blood from patients and controls was collected into PAXgene tubes (PreAnalytiX, Hombrechtikon, Switzerland) and stored at -80°C prior to RNA extraction. Total RNA was extracted from 1ml whole blood using the Preserved Blood RNA Purification Kit II (Norgen Biotek, Thorold, Canada) followed by DNAse I digestion and concentration using RNA Clean & Concentrator-5 columns (Zymo Research, Irvine, US) according to manufacturer’s instructions. All RNA samples were subsequently run on a TapeStation 2200 (Agilent, Santa Clara, US) using High Sensitivity RNA ScreenTapes. RNA integrity number (RIN) was measured from total RNA, and samples with RIN <2.5 were excluded from the study.

### Small RNA library preparation, sequencing and analysis

Small RNA libraries were prepared from total RNA using the TruSeq Small RNA Library Prep Kit (Illumina, San Diego, US) according to the manufacturer’s instructions up to and including the PCR Amplification step. Size selection was then carried out using the PippinPrep size selection system (Sage Science, Beverly, US) with 2% dye free agarose gel cassettes with Marker L, run using the program for 3% DF Marker F gel cassette. The upper and lower size selection thresholds were 137 and 200 bp respectively. The eluted size fractions were concentrated using DNA Clean and Concentrator-5 columns (Zymo Research, Irvine, US). Successful library preparation and size selection was confirmed using High Sensitivity D1000 ScreenTapes and the TapeStation 2200 (both Agilent, Santa Clara, US). Libraries were normalised to 2nM using 10mM Tris-HCL, pH8.5 and 12 libraries were pooled by combining equal volumes of each library. Illumina’s pooling guidelines were followed regarding index combinations. Pooled libraries were single end sequenced (120 cycles) with the MiSeq Reagent Kit v3 on a MiSeq sequencer (Illumina, San Diego, US) located in the University College London Genomics Laboratory (UCL Genomics) at the Institute of Child Health, London UK according to the manufacturer’s instructions. PhiX DNA was added to pooled libraries prior to sequencing to give a final concentration of 5% in order to increase the sequence diversity of the libraries.

Each MiSeq run was subjected to quality control (QC): successful runs were considered to have passed QC if the percentage of clusters passing filter, and average percentage of bases with Quality (Q) score >30/40 both exceeded 80%, and cluster density >800K/mm^2^ with even pooling. FastQ files from all MiSeq runs passing QC were downloaded from BaseSpace (Illumina, San Diego, US) and uploaded into Partek Flow (Partek Inc, St. Louis, US), along with metadata. Samples yielding less than one million reads were excluded. Adapter sequences were removed and reads trimmed to a minimum length of 15bp with a minimum quality score of 25. Reads were aligned to the human genome (hg38) using Bowtie 2 v2.2.5 with a seed length of 10 with 1 mismatch, quantified for annotated miRNAs (miRBase v21) and normalised using Cufflinks v2.2.1 default settings (total hits normalisation). Gene Specific Analysis (GSA) was carried out using a proxy for sample age, and sampling age as covariates determined by post-normalisation Principal Component Analysis (PCA), and a lowest average coverage filter of 5000 determined using a shrinkage plot.

### Relative quantification of miRNAs by reverse transcription quantitative PCR (RT-qPCR)

Power calculations based on the outcome of the small RNA-seq study determined that n=30 for both patient and control groups would be required for replication of results in an independent sample set. To this end, 10ng total RNA from all samples was reverse transcribed using the miScript II RT Kit (Qiagen, Hilden, Germany) followed by qPCR carried out using the miScript SYBR Green PCR Kit and miScript Primer assays (both Qiagen, Hilden, Germany) according to the manufacturer’s instructions. MiScript PCR Controls RNU6-2, SNORD42B, SNORD72 and SNORD95 were used as endogenous controls, and all samples were run in duplicate on a QuantStudio 12K Flex (Life Technologies, Carlsbad, US) with dissociation curves to check for specific amplification. Data was analyzed using the ΔCt method to generate ratios of expression of test miRNA compared to endogenous controls, and the ΔΔCt method to generate fold changes between patient and control groups.

### miRNA target identification and expression

Target genes for hsa-miR-16-5p, hsa-miR-93-5p and hsa-let-7i-5p were selected using TarBase v8 from DIANA tools^56^. Genes validated by luciferase assay were prioritized. Where no luciferase data was available (hsa-let-7i-5p), targets validated by immunoprecipitation in >5 publications were selected. Shortlisted targets for each miRNA with the highest level of expression in whole blood (GTEx, https://gtexportal.org/home/)^53^ were then selected for expression analysis in the replication cohort (Supplementary Table 3).

cDNA preparation and qPCR analysis were performed as previously described^57^. Each experiment was performed in at least two technical replicates on a QuantStudio 12K Flex. Primers were purchased from Qiagen for *CCND3* (hsa-miR-16-5p target), *VEGFA* (hsa-miR-16-5p and hsa-miR-93-5p target), *CDKN1A* (hsa-miR-93-5p target), *ZFP36, NAP1L1* and *RNF44* (hsa-let-7i-5p targets), *B2M, ALAS1* and *ACTB* (endogenous controls), and *MCM7*. Qiagen primer reference numbers were as follows: *CCND3* QT00096796; *VEGFA* QT01010184; *CDKN1A* QT00062090; *ZFP36* QT00091357; *NAP1L1* QT00085635; *RNF44* QT00051793; *MCM7* QT00053228; *ACTB* QT01680476; *B2M* QT00088935; *ALAS1* QT00073122. Gene set enrichment analysis was performed using the ENRICHR program (http://amp.pharm.mssm.edu/Enrichr/)^58,59^.

### ROC analysis

ROC curves were plotted, and AUCs calculated, in SPSS v26 (IBM, Armonk, US) using Z scores calculated from either log transformed normalised small RNA-seq read counts, or from log transformed qPCR ratios of miRNA expression measured relative to endogenous control miRNAs. For combined analysis of hsa-let-7i-5p, hsa-miR-16-5p and hsa-miR-93-5p, mean Z scores for these miRNAs were calculated and used to plot the ROC curve. The sCJD vs AD comparison was carried out indirectly, using qPCR data from each group obtained using a common set of control samples (Figs. 2a, 3, and Supplementary Figs. 3a-d and 5a-d). Youden’s indices were calculated using ROC curve coordinates, in order to determine sensitivity and specificity at the point where test performance is optimal.

### Statistical analysis

Small RNA-seq differential expression p values were adjusted for multiple testing using the Benjamini-Hochberg false discovery rate (FDR) method. miRNA expression data from the discovery cohort sCJD group was subjected to Pearson or Spearman correlation (as appropriate) with clinical parameters after first checking data for normality. Clinical parameters tested were age of onset, duration of disease, MRC Scale score and MRC Scale slope, and correlation p values were Bonferroni corrected for multiple testing. For miRNA and mRNA qPCR data, statistical comparisons between groups were carried out using Prism v5.03 (GraphPad, San Diego, US) by two-tailed Mann-Whitney test, or one-tailed Mann-Whitney test where there was prior knowledge of the direction of effect e.g. for qPCR replication of discovery cohort data. Longitudinal data was analyzed by linear regression using Prism.

### Data Availability

All sequencing data that support the findings of this study have been deposited in the National Center for Biotechnology Information Gene Expression Omnibus (GEO) and are accessible through the GEO Series accession number GSE140069. The source data underlying Table 1, Figures 1a,b, and 2-5, and Supplementary Figures 2-11 are provided as a Source Data File.

## Data Availability

The sequencing data discussed in this publication have been deposited in NCBI's Gene Expression Omnibus (Edgar et al., 2002) and are accessible through GEO Series accession number GSE140069.

https://www.ncbi.nlm.nih.gov/geo/query/acc.cgi?acc=GSE140069

## Acknowledgements

We are very grateful to patients and volunteers who have made this study possible by donating blood samples and their time to research; Holger Hummerich for helpful discussions; Tony Brooks, Paola Niola, and Mark Kristiansen from UCL Genomics for technical assistance with MiSeq; Tracy Campbell and Joanna Field for help with metadata acquisition; Richard Newton for help with preparation of figures.

## Author Contributions

S.M. had full access to all data in the study and takes responsibility for integrity of the data and accuracy of the data analysis. All authors read and approved the final manuscript. Concept and design: E.A.V, S.M; Acquisition, analysis, or interpretation of data: all authors; Drafting of manuscript: P.J.N, E.A.V, S.M; Statistical analysis: P.J.N; Obtaining funding: J.C, S.M; Administrative, technical, or material support: P.J.N, E.A.V, S.M, J.C; Study supervision: S.M, E.A.V.

## Materials and Correspondence

Correspondence and material requests to be addressed to S.M.

